# The brief comparison of the operational efficiency of pool-testing strategies for COVID-19 mass testing in PCR laboratories

**DOI:** 10.1101/2020.07.14.20151415

**Authors:** Kirill Vechera

## Abstract

This paper addresses the operational efficiency of different pool-testing strategies in typical scenarios of a PCR laboratory working in mass testing for COVID-19 with different values of prevalence, limitations and conditions of testing, and priorities of optimization.

The research employs a model of the laboratory’s testing process, created after interviewing of PCR laboratories and studying their operations. The limitations and operational characteristics of this model were applied in a simulation of the testing process with different pool-testing strategies managed by a computer program developed in the LOMT project.

The efficiency indicators assessed are the number of assays needed to obtain results of a batch of specimens, the number of specimens identified after the first analysis, and total time to obtain all results.

Depending on prevalence, constraints of testing, and priorities of optimization, different pool-testing strategies provide the best operational efficiency. The binary splitting algorithm provides the maximum reduction of the number of assays: from 1.99x reduction for a high prevalence (10%) to 25x reduction for a low prevalence (0.1%), while other algorithms provide the least amount of time to obtain results or the maximum number of the specimens classified after the first analysis.

## Introduction

For a PCR laboratory, the major indicator of the efficiency of a pool-testing strategy is the reduction of the number of assays for a certain number of specimens. The analysis process takes up to 4 hours, consumes reagents and requires significant manual labor. Other related processes require much less time and cost, and we can assume that a tenfold reduction in the number of assays increases the throughput of the laboratory by 9 times and also decreases the cost of the testing of one person by 9 times.

The number of assays during pool testing primarily depends on prevalence — that is, the proportion of positives samples. The lower the prevalence, the larger the pool size that can be used, and ultimately, fewer assays will be required. For example, for 1% prevalence, the maximal effective pool size is 69, for 10% prevalence it is 7 and for prevalence above 25% the pool size becomes less than 4 and the efficiency becomes too low to be worth using.

For a PCR laboratory, two other factors are important: the limit of dilution of a specimen and the time it takes to get results for a full batch of specimens.

The limit of dilution here is the maximum number of specimens that are mixed in a single pool, or the maximum pool size. The limit of dilution depends on the sensitivity of a test and its limit of detection, since if the dilution of a positive specimen leads to the amount of its detecting material decreasing below the limit of detection, the specimen is erroneously defined as negative. Studies have shown that a pool size of up to 32 allows for the confident identification of asymptomatic people in RT-PCR testing for SARS-CoV-2 RNA ^1, 2^.

The processing time of a batch of specimens depends not only on the number of specimens and how fast the laboratory can perform analyses but also on the depth (the number of stages) of the pooling strategy. Any pooling strategy requires two or more stages to obtain results for all the specimens in a batch, and each stage can begin only after the previous stage has been completed. In most cases, the more stages the pooling strategy has, the more assays are saved, but also the more time is required to get the results of a full batch.

As analyses are performed, some specimens are identified as positive or negative from the analysis results, while other specimens remain for repeated analyses in the next stages. The number of samples identified after the first analysis can vary significantly with different strategies. Algorithms in which one sample goes into only one pool at each stage can identify positive samples only at the last stage, while all other stages identify negative samples only. Combinatorial algorithms in which one sample goes into several pools are capable of identifying both negative and positive samples at any stage and can also obtain the results of a full batch in many cases in the single stage, requiring the second stage for only some rare cases.

There are also non-adaptive algorithms that the result of all samples to be obtained in one stage, at the cost of reducing the accuracy of detection and producing a certain number of false positives. While it can be useful in some other tasks, we see no sense in using such algorithms for COVID-19 testing of people, so these algorithms are not considered here.

## Methodology

For the research, 11 PCR laboratories in Italy, Russia and the USA working in COVID-19 mass testing were interviewed for the assessment of the throughput, time, and constraints of individual operations within the laboratory. Pool-testing strategies were executed with the computer program for managing pool-testing workflows, which was developed in our project LOMT - Laboratory Optimizer for Mass Testing ^3^. The efficiency indicators ^4^ were calculated in a simulation of the laboratory testing process with a pool-testing workflow managed by the LOMT program. Three datasets with 1000 batches of 1000 specimens were created from random data with positive element probabilities *p* = 0.1, *p* = 0.01, and *p* = 0.001 with normal distribution.

## The comparison

Different laboratories can operate with different values of prevalence, constraints of testing and priorities of optimization, and these values can change over time. LOMT supports several different pooling strategies and automatically searches for the optimal strategy for every specific condition. This comparison provides an assessment of the efficiency of the most used strategies in several typical scenarios.

The task consists of RT-PCR testing of 1000 swabs on a 1 thermal cycler with 96 wells; full analysis time is 4 hours, the dilution limit is 32, and high, medium and low prevalence rates are 10%, 1% and 0.1%, respectively. The time to transfer fluid from specimen tubes to assay tubes is not taken into account, although it can slightly increase using combinatorial algorithms.

The efficiency indicators assessed are as follows:

1. The number of assays — the smaller the number, the more laboratory throughput increases and the cost of testing one person decreases.
2. First results — the number of specimens completely classified after the first analysis (after the shortest possible time) and the number of positive specimens found.
3. Total time to result — the time (hours) required to obtain the results of all swabs in the batch; some swabs from the batch are mixed in pools and analyzed the entire time.

### Individual

In the standard testing method, without pooling, all specimens are tested individually. We use this as a baseline to assess the efficiency of pool-testing strategies. A prevalence rate of 10% means there are 900 negative specimens and 100 positive ones; a prevalence rate of 1% means 990 negative specimens and 10 positive ones; and a prevalence rate of 0.1% means 999 negative specimens and 1 positive.

| Prevalence | 10% | 1% | 0.1% |
| --- | --- | --- | --- |
| Assays | 1000 | 1000 | 1000 |
| First results | 94 | 94 | 94 |
| First results, positives | 9.4 | 0.94 | 0.094 |
| Total time to result, h | 44 | 44 | 44 |

The efficiency is the same for any prevalence from 0 to 100% — 1000 swabs need 1000 assays. First results are available in 4 hours — these are 94 swabs with the number of positive samples proportional to the prevalence of the batch. Total time to obtain results is 44 hours, with the manual labor and thermal cycler operations as the bottleneck.

### Plain

This is a classic strategy of pool testing, which runs in two stages: in the first stage, all the specimens are split into several groups, and each group is mixed into a single pool and tested. A negative result means that all the specimens in the pool are negative. The pools with a positive result go to the second stage, in which all specimens are tested individually.

|  |  |  |  |
| --- | --- | --- | --- |
| **Prevalence** | 10% (pool 4) | 1% (pool 10) | 0.1% (pool 32) |
| **Assays** | 594 (1.68) | 196 (5.1) | 63 (15.9) |
| **First results** | 246 (2.62) | 895 (9.5) | 969 (10.3) |
| **First results, positives** | 0 | 0 | 0 |
| **Total time to result, h** | 28 (1.57) | 12 (3.67) | 8 (5.5) |

With 10% prevalence, 1000 specimens require 594 assays (1.68x reduction compared to 1000 assays at baseline). First results are available in 4 hours; 250 specimens, only negative ones (2.64x increase compared to 94 at baseline). Total time to obtain results is 36 hours (1.22x reduction compared to 44 hours at baseline).

With 1% prevalence, 1000 specimens require 196 assays (5.1x reduction). First results: 846 specimens, only negative ones (9x increase). Total time to obtain results is 16 hours (2.75x reduction).

With 0.1% prevalence, 1000 specimens require 63 assays (15.9x reduction). First results: 970 specimens, only negative ones (10.3x increase). Total time to obtain results is 8 hours (5.5x reduction).

### Plain Multilevel — 2 levels

This strategy is a classic pool-testing strategy with several pooling stages instead of a single one. The pools with positive results go to the next stage, and the pool size decreases with each stage. This approach significantly reduces the number of samples that are tested individually at the last stage compared to the single-level pooling. The higher the number of levels, the fewer assays needed. The values are for the strategy with two levels of pooling.

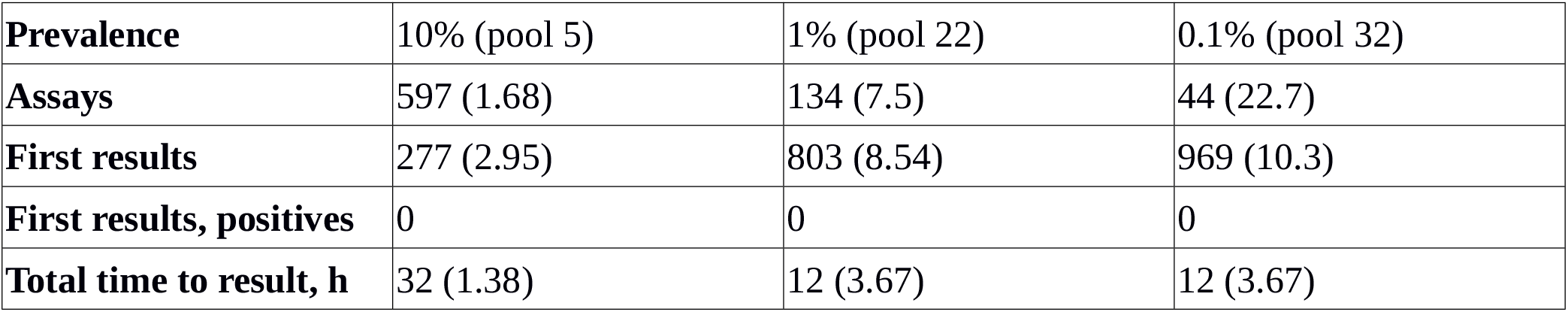

|  |  |  |  |
| --- | --- | --- | --- |
| **Prevalence** | 10% (pool 5) | 1% (pool 22) | 0.1% (pool 32) |
| **Assays** | 597 (1.68) | 134 (7.5) | 44 (22.7) |
| **First results** | 277 (2.95) | 803 (8.54) | 969 (10.3) |
| **First results, positives** | 0 | 0 | 0 |
| **Total time to result, h** | 32 (1.38) | 12 (3.67) | 12 (3.67) |

### Bisector Waiting

This strategy applies the parallelized version of Generalized Binary Splitting algorithm. It provides the maximum possible reduction of the number of assays but increases the pooling depth incrementing total time to result. For 10% prevalence it is 13 stages on average, for 1% prevalence it is 9 stages and for 0.01% it is 5 stages.

| Prevalence | 10% (pool 8) | 1% (pool 32) | 0.1% (pool 32) |
| --- | --- | --- | --- |
| Assays | 477 (2.1) | 90 (11.1) | 39 (25.6) |
| First results | 323 (3.43) | 730 (7.77) | 969 (10.3) |
| First results, positives | 0 | 0 | 0 |
| Total time to result, h | 56 (0.79) | 36 (1.22) | 20 (2.2) |

### Bisector Waiting with a depth limit

This strategy limits the pooling depth by executing individual testing of all specimens left after the depth limit is reached. The number of specimens in the last stages is small, so switching to the individual testing slightly decreases the assays savings but improves the batch processing time.

| Prevalence | 10% (pool 7, depth 7) | 1% (pool 32, depth 7) | 0.1% (pool 32, depth 4) |
| --- | --- | --- | --- |
| Assays | 503 (1.99) | 100 (10) | 47 (21.3) |
| First results | 314 (3.34) | 730 (7.77) | 969 (10.3) |
| First results, positives | 0 | 0 | 0 |
| Total time to result, h | 36 (1.22) | 28 (1.57) | 12 (3.66) |

### Enumerator Doublets

This strategy uses a combinatorial algorithm in which every specimen goes into two different pools. This approach allows for the classification of the maximum number of specimens at the first stage, and it detects both negative and positives swabs. The efficiency of combinatorial strategies increases with decreasing prevalence. A simplified variant of the strategy is called also Array testing or Matrix testing.

| Prevalence | 10% (pool 8, depth 3) | 1% (pool 32, depth 3) | 0.1% (pool 32, depth 2) |
| --- | --- | --- | --- |
| Assays | 562 (1.78) | 118 (8.5) | 65 (15.4) |
| First results | 247 (2.91) | 920 (9.8) | 998 (10.6) |
| First results, positives | 0.67 | 0.01 | 0.56 |
| Total time to result, h | 28 (1.57) | 12 (3.67) | 8 (5.5) |

### Enumerator Triplets

This is similar to the previous strategy with a combinatorial algorithm, in which every specimen goes into three pools. The strategy is able to identify more positive swabs in the first stages. For 0.1% prevalence, this strategy gives results for all specimens immediately after the first assay in 93% of cases, and requires a second stage of assays only in the remaining 7% cases.

|  |  |  |  |
| --- | --- | --- | --- |
| **Prevalence** | 10% (pool 8, depth 5) | 1% (pool 32, depth 3) | 0.1% (pool 32, depth 2) |
| **Assays** | 595 (1.68) | 125 (8) | 95 (10.5) |
| **First results** | 185 (1.96) | 954 (10.1) | 999 (10.6) |
| **First results, positives** | 5.8 | 3 | 0.9 |
| **Total time to result, h** | 36 (1.22) | 12 (3.67) | 8 (5.5) |

## Summary

After comparing these strategies, we can see which are best for the maximum increment of the throughput of a laboratory (by reducing the number of assays) and which are best for reducing total time to process a batch of swabs.

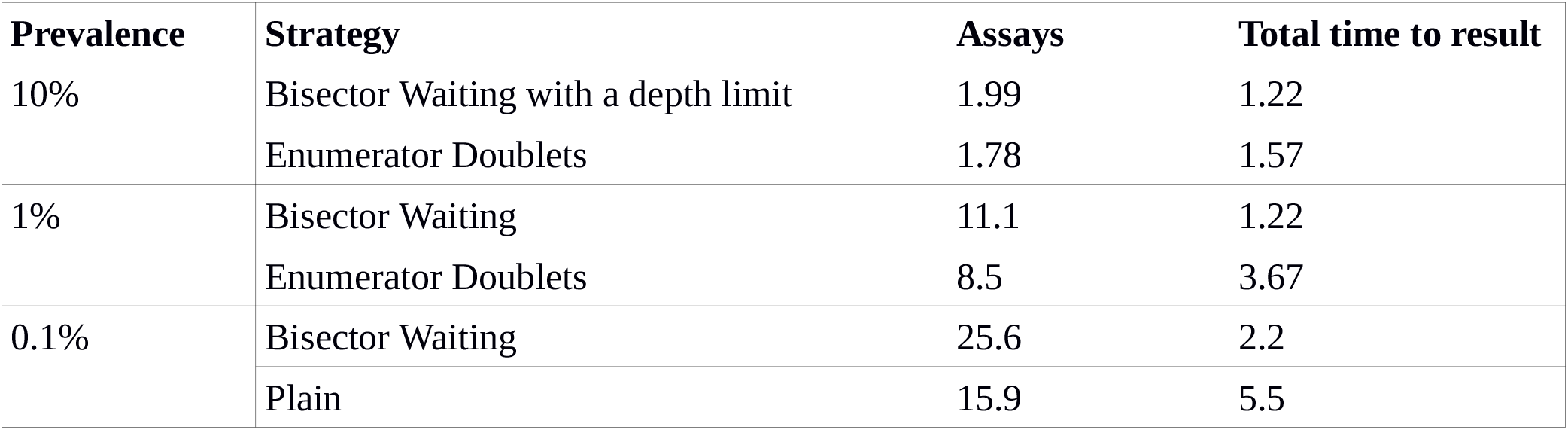
The reduction of assays and total time to obtain results for different prevalence values and pool testing strategies

For a high prevalence (10%), the Bisector Waiting strategy with a depth limit gives the best assay savings and throughput increment by **1**.**99** times, while the Enumerator Doublets strategy gives the best assay savings by **1**.**78** times with the least amount of time to obtain results — **1**.**57** times less compared to the individual testing.

For a medium prevalence (1%), the Bisector Waiting strategy gives the best assay savings and throughput increment by **11**.**1** times, and the Enumerator Doublets strategy gives the best assay savings by **8**.**5** times with the least amount of time to obtain results — **3**.**67** times less compared to the individual testing.

For a low prevalence (0.01%), the Bisector Waiting strategy again gives the best assay savings and throughput increment by **25**.**6** times, while the Plain strategy gives the best assay savings by **15**.**9** times with the least amount of time to obtain results — **5**.**5** times less compared to the individual testing.

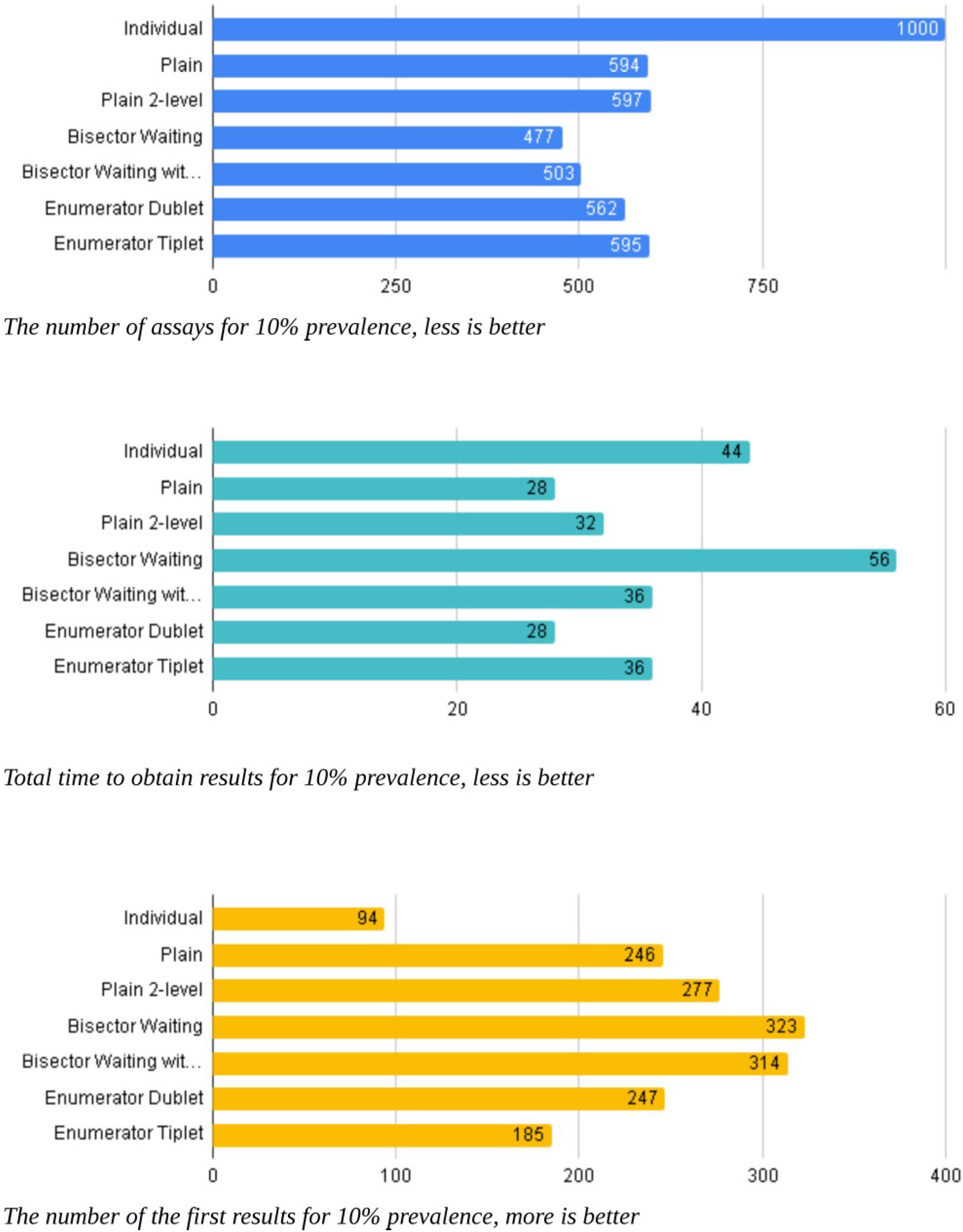

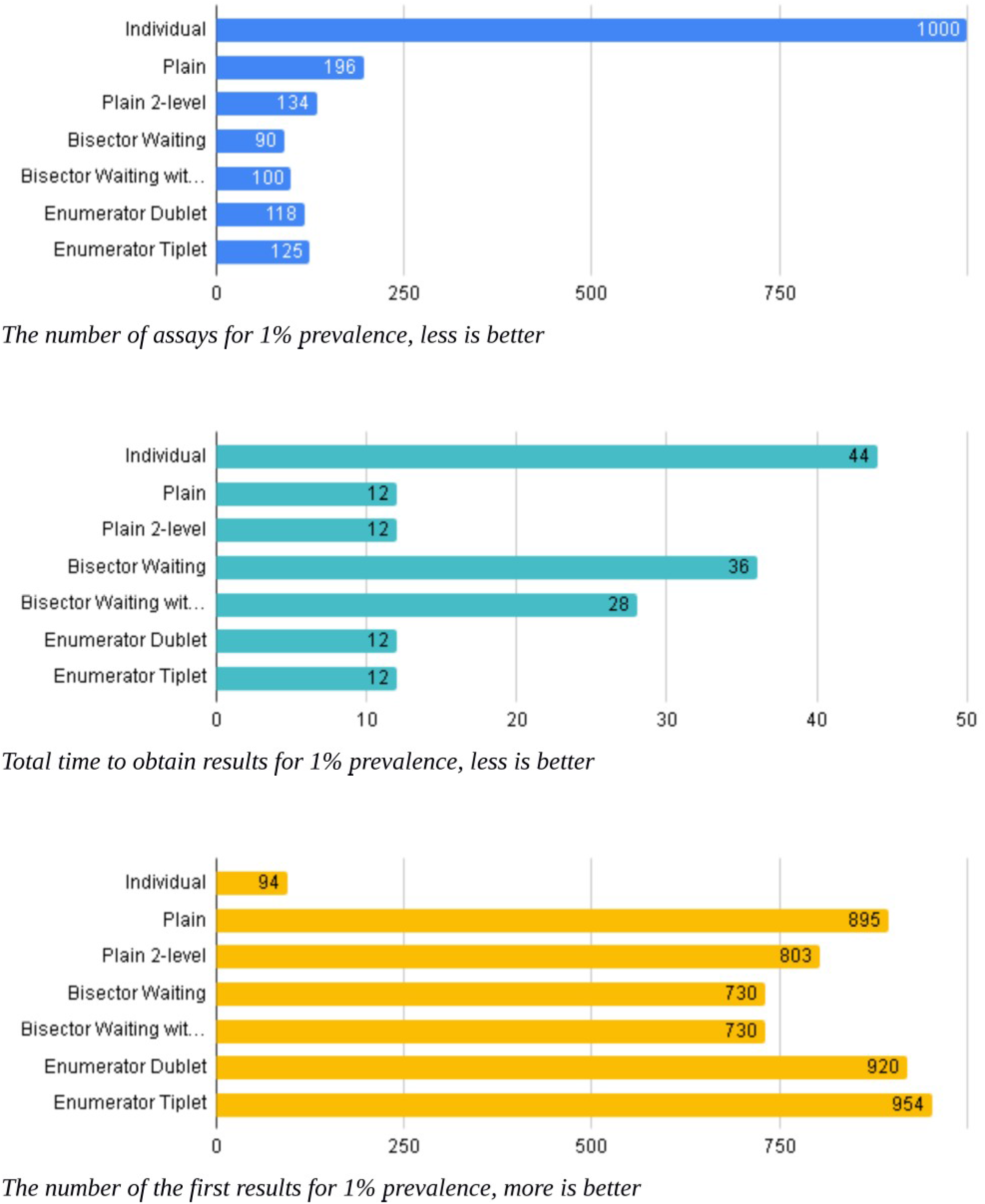

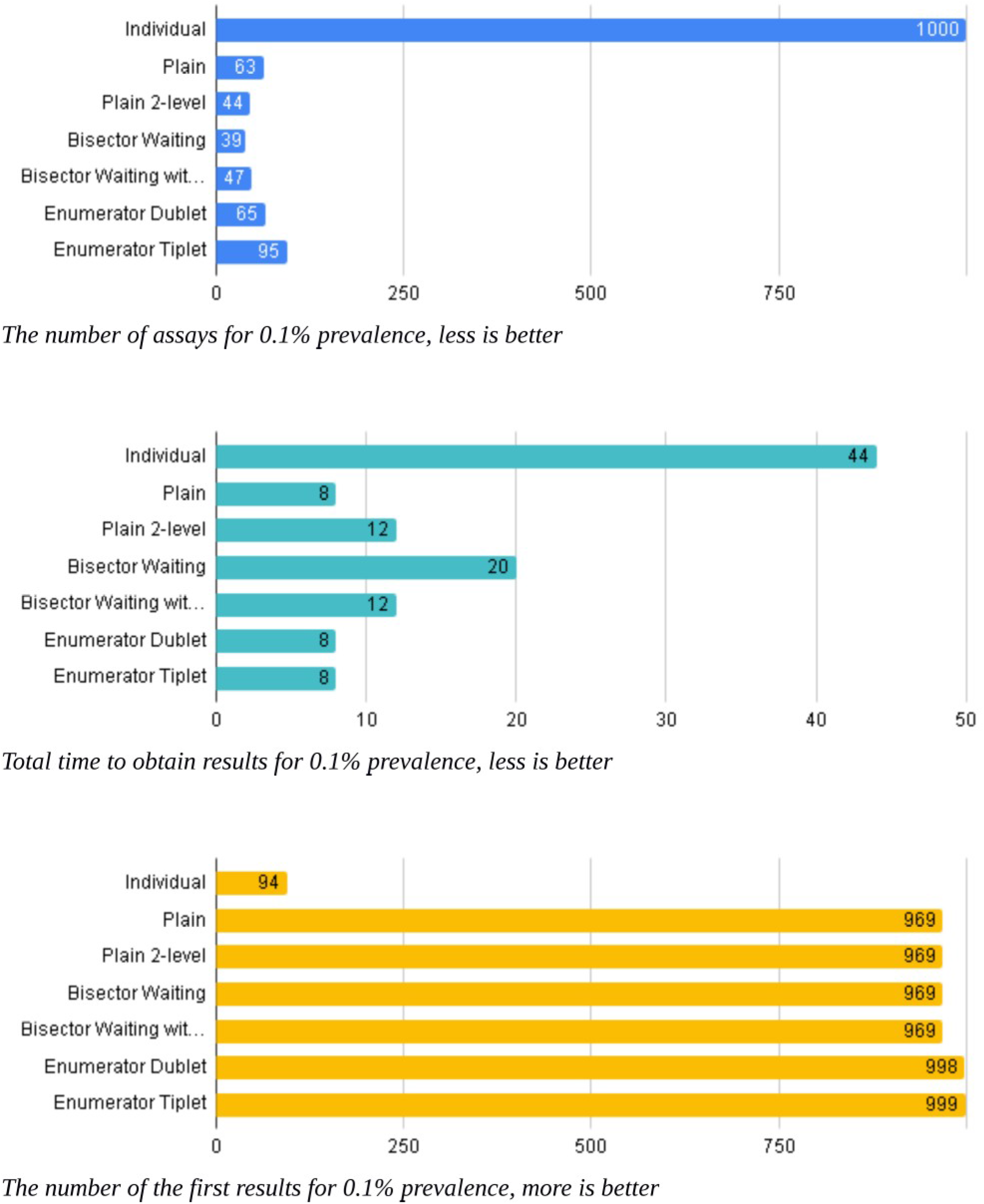

## Data Availability

All data and code is available here at the link provided: https://lomt.jetware.org/download/articles/operational_efficiency_pool_testing-3.tar.xz

## Funding

European Open Science Cloud, COVID-19 Fast Track Funding

## Notes

### Competing Interest Statement

The authors have declared no competing interest.

### Author Declarations

All relevant ethical guidelines have been followed

